# The efficacy and safety of remdesivir in the treatment of patients with COVID-19: a systematic review and meta-analysis

**DOI:** 10.1101/2021.03.12.21253470

**Authors:** Lixiang Lou, Hui Zhang, Zeqing Li, Baoming Tang, Zhaowei Li

## Abstract

**Background:** The global total of COVID-19 cases will reach 20 million this week, with 750,000 deaths. It has spread to more than 200 countries and regions around the world. At present, the global pandemic continues to rise and continues to spread worldwide. It is necessary to explore the effective and safe treatment of COVID-19 as soon as possible. Remdesiviras was an antiviral agent with therapeutic potential, but it was still controversial.

**Objective:** Through systematic review and meta-analysis, to evaluate the effect and safety of remdesivir in the treatment of patients with COVID-19, and will provide a reliable reference for the treatment of COVID-19.

**Methods:** We used the following search string: “COVID-19” [Mesh], “remdesivir” [Mesh], “randomized controlled trial” [Mesh]. We used the Medical Subject Heading (MeSH) terms and corresponding keywords to make the search strategy. We searched six databases, PubMed, EMBASE, Cochrane Library, Web of Science, clinical trials.gov and chictr.org.cn. Data analyses were conducted by using the software Review Manager 5.3 and STATA version 14.0.

**Results:** Our systematic search identified 5 meta-analyses of RCTs, including 1782 patients with COVID-19.The clinical improvement of remdesivir in the treatment of COVID-19 was superior to the placebo-controlled group (relative risk (RR) =1.17, 95% confidence interval (CI)=1.07-1.29, *p*=0.0009). The following are the Single-Arm Study, Meta-analysis results. The pooled prevalence of clinical improvement significant findings was 62% (95% CI = 59-65%, *p*=0.00), during treatment of COVID-19 with remdesivir. The incidence rates of Acute kidney injury, Hepatic enzyme increased, Any serious adverse event were 5% (95%CI=3-7%, *p*=0.00), 11%(95%CI=5-16%, *p*=0.00), 22%(95%CI=18-27%, *p*=0.00), respectively, and the mortality was 13%(95%CI=8-19%, *p*=0.00), during treatment of COVID-19 with remdesivir.

**Conclusion:** This analysis confirms that remdesivir is effective in the clinical improvement of COVID-19 patients, and the rate of clinical improvement was 62%. In addition, adverse events and mortality should also be paid attention to. Future research should aim that more large-scale studies were needed to confirm the results, to further elucidate the underlying mechanisms.

## Introduction

Coronavirus disease 2019 (COVID-19) is caused by a novel coronavirus called SARS-CoV-2. SARS-CoV-2 shares 79% RNA sequence identity with acute respiratory syndrome coronavirus (SARS-CoV), and 50% genomic sequence with Middle East respiratory syndrome coronavirus (MERS-CoV) [1–4]. It is an emerging human infectious coronavirus originally reported in Wuhan’s seafood market, and spread rapidly around the world in a short time. The global total of COVID-19 cases will reach 20 million this week, with 750,000 deaths. It has spread to more than 200 countries and regions around the world. At present, the global pandemic continues to rise and continues to spread worldwide. On March 11th, 2020, the World Health Organization (WHO) has declared a global pandemic [5]. Therefore, it is necessary to explore the effective and safe treatment of COVID-19 as soon as possible. remdesiviras was an antiviral agent with therapeutic potential, and Antiviral Activity against RNA Viruses was been confirmed by research [6,7]. Remdesivir (GS-5734), an inhibitor of viral RNA-dependent RNA polymerase, has inhibitory activity against SARS-CoV and Middle East Respiratory Syndrome (MERS-CoV) [8–11] and was identified as very early a potential therapeutic candidate for COVID-19,because of its ability to inhibit SARS-CoV-2 in vitro [12]. Preliminary evidence from in vitro and animal studies is provided to demonstrate the clinical therapeutic potential of remdesivir, for human infections caused by contemporary and emerging coronaviruses (including SARS-CoV-2) [13]. The U.S. Food and Drug Administration has issued an Emergency Use Authorization (EUA) to allow the use of remdesivir to treat adults and children hospitalized for severe coronavirus disease 2019 (COVID-19), but the results are still controversial. Therefore, we conducted a systematic review and meta-analysis based on existing data,to analyze the efficacy and safety of remdesivir in the treatment of patients with COVID-19, and provide a reference for clinical treatment.

## 1 Methods

### 1.1 Retrieval of published studies

Adhering to PRISMA guidelines, a literature search was conducted on June 13, 2020, using the following search string: “COVID-19” [Mesh], “remdesivir” [Mesh], “randomized controlled trial” [Mesh]. We used the Medical Subject Heading (MeSH) terms and corresponding keywords to make the search strategy. We searched six databases, PubMed, EMBASE, Cochrane Library, Web of Science, clinical trials.gov, and chictr.org.cn. The search was restricted to the English language. Two reviewers (LL and BT) independently screened the literature search and assessed each study for inclusion. The possible disagreement between reviewers was resolved by consulting a senior investigator (ZL).

### 1.2 Inclusion and exclusion criteria

#### 1.2.1 Inclusion criteria

(1)Methods Study Design and Participants remdesivir trial is a randomized controlled trial (RCT). (2)Blood or respiratory samples were positive by real-time polymerase chain reaction test for the nucleic acid of COVID-19. (3)The experimental group (conventional treatment combined with remdesivir) and the control group (given placebo) were treated under the same conditions as conventional treatment, or the intervention group, the experimental group and the control group, were treated with remdesivir treatment. (4)Mainly compare the effective number of the experimental group, and the control group (clinical improvement, liver transaminase increase, acute kidney injury, serious adverse events, the number of deaths.

#### 1.2.2 Exclusion criteria

(1)Animal experiment. (2)review papers, commentary articles, publication in other languages, duplicated reports and case series, and the full text cannot be obtained. (3)The outcomes were unclear.

### 1.3 Data extraction

Two of us (LL, HZ) undertook data extraction independently, using a previous pilot data extraction form. Any disagreement was resolved by discussion between the two reviewers. A customized form was used to record the authors of the study, publication time, design of the trial, country or region, the number of COVID-19 patients, patient age, sex, time from the onset of symptoms to hospitalization (days), clinical improvement, Hepatic enzyme increased, acute kidney injury, any severe adverse event, Death. Second, we also screened them according to that blood or respiratory samples were positive by real-time polymerase chain reaction test for the nucleic acid of COVID-19. In addition, we also conducted a one-arm meta-analysis for data. We have analyzed the prevalence of clinical symptoms among COVID-19 patients used by remdesivir. A study contained two arms, the data were abstracted separately for each comparator.

### 1.4 Quality assessment of included meta-analyses

The quality of studies was assessed based on randomization, masking, and withdrawal. The quality of eligible meta-analyses was assessed using Cochrane systematic review bias risk assessment tool. In addition, a modified JADAD ranking scale was used to assess the quality of randomized, controlled trials.

### 1.5 Statistical analysis

Data analyses were conducted by using the software Review Manager 5.3 and STATA version 14.0. we estimated the effect of remdesivir on COVID-19. Heterogeneity was quantified using the I-squared (*I*^*2*^) statistic, and categorized as low (*I*^*2*^ <25%), moderate (*I*^*2*^ =25-50%) or high (*I*^*2*^ >50%). A model of fixed effect (FE) was set with 95% CI if no statistical evidence of heterogeneity existed (*p*≥ 0.1, *I*^*2*^ ≤ 50%), while a model of random effect (RE) was applied to estimate pooled effect with 95% confidence intervals (CI) if statistical heterogeneity was found (*p* < 0.1, *I*^*2*^ > 50%). If significant heterogeneity (*p*< 0.1, *I*^*2*^ > 75%) was founded, we used sensitivity analysis to explore its possible sources. A random-effects model was used if the reason for heterogeneity could not be identified. We visually inspected the Begg’s funnel plot asymmetry. Meanwhile, we applied Begg’s test to evaluate publication bias. Probability (*p*) values less than 0.05 were considered statistically significant.

## 2 Results

### 2.1 Systematic search results

The literature search based on the search strategies produced 310 citations, including 166 from PubMed, 48 from Embase, 13 from Cochrane, 81 from Web of Science, 2 from clinical trials.gov. Search results from different sources were entered into an EndNote library, which was reduced to 240 after duplicates were removed. According to the title and the abstract screening removed 138 articles, leaving 33 items. After reading the remaining 33 full-text articles, 27 studies were removed for at least one of following reasons:(1)Non-randomized controlled trials or The research Method inconsistency; (2) Outcome indicators are inconsistent; (3)Results repeated publication. Ultimately, 5 studies [14–18] were included in the meta-analysis involving 1780 participants, of which were RCTs (Figure 1).

**Figure 1.**
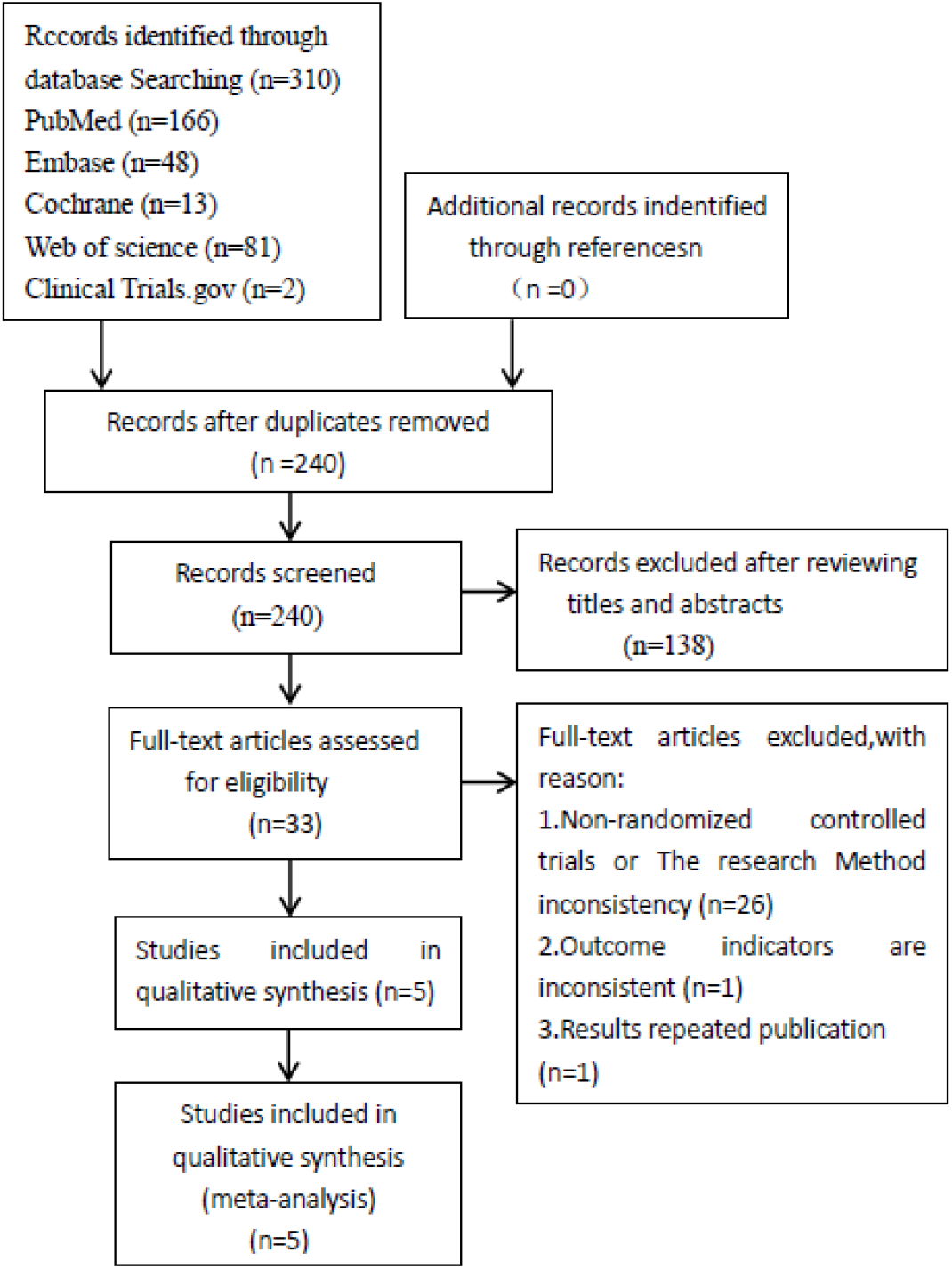
Flow chart of literature search and selection of studies. A total of 310 records were initially identified. The remaining five RCTs were included in the final meta-analysis.

### 2.2 Characteristics of trials

The RCTs included were mainly undertaken in the 15 countries or regions. Two articles [15,18] were randomized placebo-controlled trials, and there were 868 patients in the remdesivir groups and 846 in the placebo-controlled groups. One study [18] was terminated from the study before its completion, because there has been a decrease in COVID-19 cases in China. Moreover, we conducted a single-arm study analysis according to different outcomes. Five studies [14–18] described clinical improvement, and there were a total of 1181 patients, with 731 events. Four studies [14, 16–18] described Elevated liver transaminase, and there were a total of 643 patients, with 69 events. Four studies [14, 16–18] described acute kidney injury, and there were a total of 643 patients, with 27 events. Five studies [14–18] described Adverse severe events, and there were a total of 1181 patients, with 284 events. Five studies [14–18] described Death, and there were a total of 1181 patients, with 107 events (Table 1).

**Table 1.**
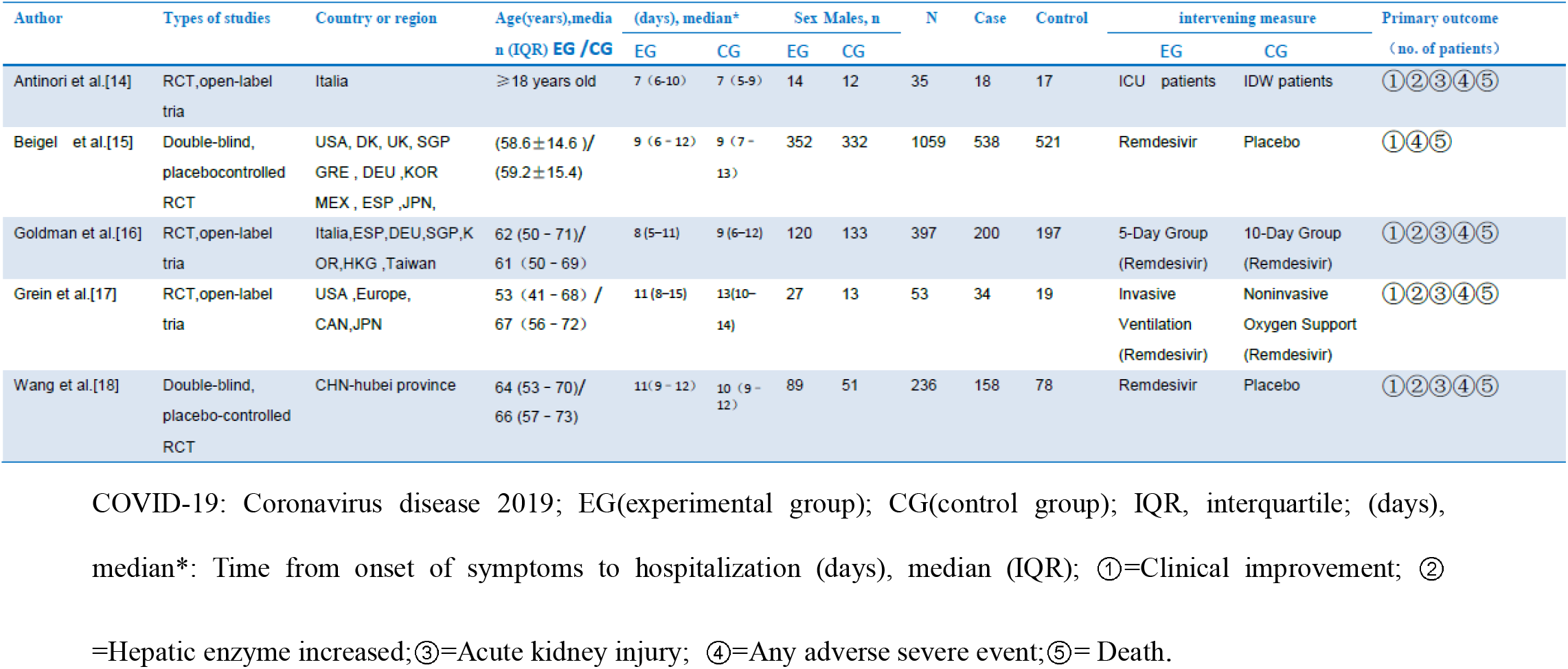
Characteristics of studies included in the meta-analysis. The detailed characteristics of the 5 studies included in this meta-analysis are provided in Table 1.

### 2.3 Methodological assessment of included Literature

The Cochrane Risk of Bias Tool was used to assess study quality. As Figure 2 shows, one study showed a low risk of bias. In addition, according to the Jadad scale score, 3 of the five documents included were high-quality studies, and 2 were low-quality studies, resulting in low-quality research (Table 2). It may be difficult to achieve blindness because remdesivir is a clinical three-phase open experiment.

**Table 2.** Methodology –Quality Assessment-Modified Jadad Score (7-point). 3 of the five documents included were high-quality studies, and 2 were low-quality studies, resulting in low-quality research (Table 2).

| Study |  | randomization | concealment<br>of allocation | double-<br>blinding | Withdrawals<br>and dropouts | Total<br>score |
| --- | --- | --- | --- | --- | --- | --- |
| Antinori et al | 2020 | 1 | 1 | 0 | 1 | 3 |
| Beigel et al | 2020 | 2 | 1 | 1 | 1 | 5 |
| Goldman et al | 2020 | 1 | 1 | 0 | 1 | 3 |
| Grein et al | 2020 | 2 | 1 | 0 | 1 | 4 |
| Wang et al | 2020 | 2 | 2 | 1 | 1 | 6 |

**Figure 2.**
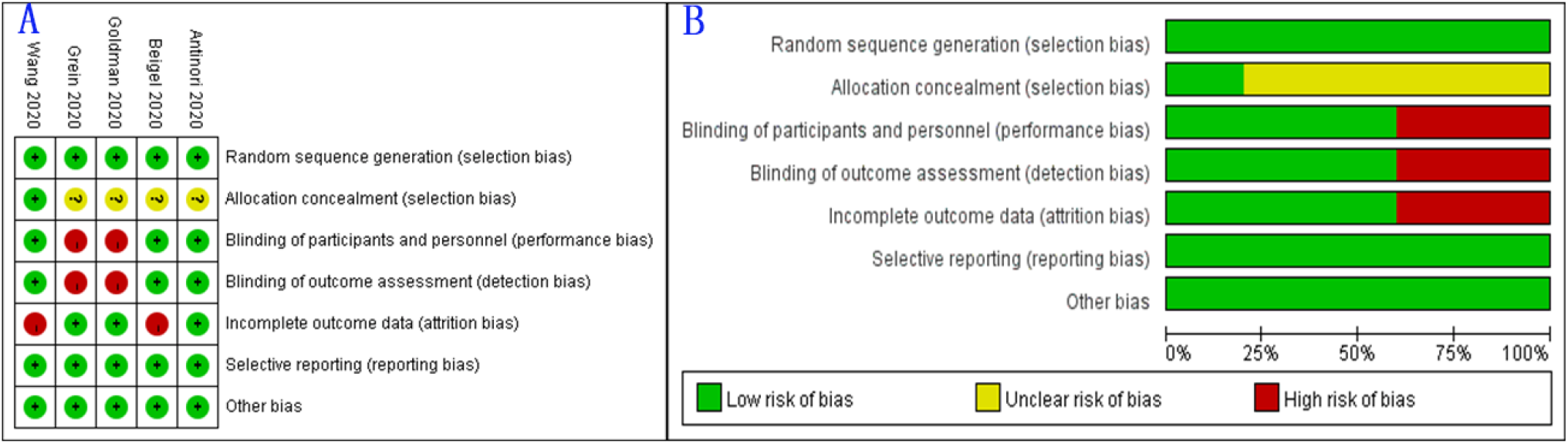
A: Cochrane systematic review bias risk assessment tool; B: Risk assessment chart. As Figure 2 shows, one study showed a low risk of bias.

### 2.4 Placebo-controlled RCT, Meta-analysis results

#### 2.4.1 Heterogeneity test

For two articles[15,18], Q-test *p*=0.7 > 0.1 and heterogeneity statistic *I*^*2*^=0%< 25% for heterogeneity between studies, suggesting no statistically significant between-study heterogeneity. A fixed-effects model was applied to estimate pooled relative risk (RR).

#### 2.4.2 The meta-analysis of fixed-effects model

Pooled RRs from two studies showed statistically significant in clinical improvement (RR□=□1.17, 95% CI (1.07-1.29), suggesting that the clinical improvement of remdesivir in the treatment of COVID-19 is better than the placebo-controlled group. Moreover, There was a marked, statistically significant between the remdesivir group and the placebo control group (Z=3.33, *p*=0.0009<0.05). Figure 3 presents the forest plot of included studies.

**Figure 3.**
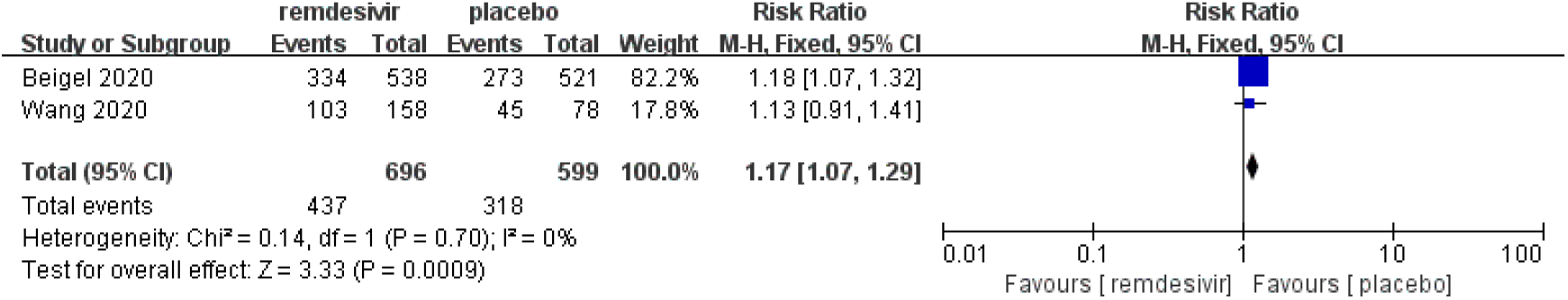
Forest figure: placebo-controlled RCT. Figure 3 presents the forest plot of included studies.

#### 2.4.3 Publication Bias

The potential publication bias was assessed by funnel plot and Begg’s bias test. The shape of the funnel plot was almost symmetrical, suggesting that there was no publication bias (Figure 4). In addition, the results of Begg’s Test (*p* □ = □1.00) was indicated no evidence of statistically significant publication bias among the included studies.

**Figure 4.**
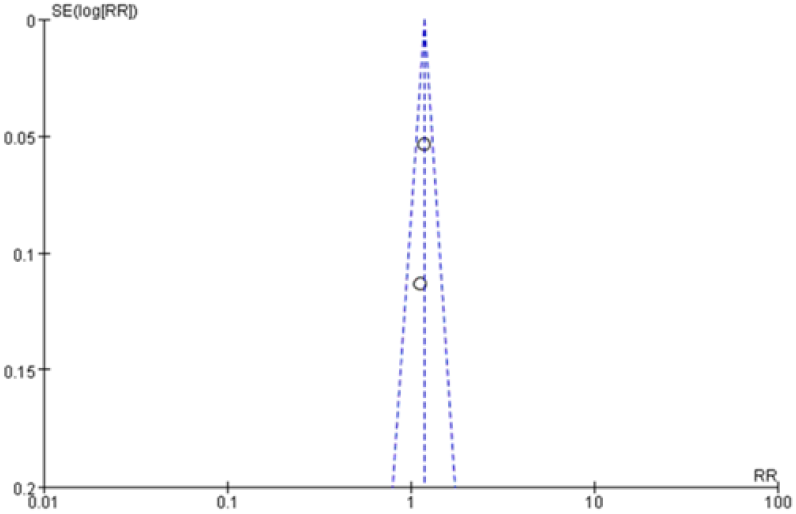
Funnel plot: placebo-controlled RCT. The shape of the funnel plot was almost symmetrical, suggesting that there was no publication bias.

### 2.5 Single-Arm Study: Clinical improvement, Meta-analysis results

#### 2.5.1 Heterogeneity test

In these five articles [14–18], Q-test *p*=0.65 > 0.1 and heterogeneity statistic *I*^*2*^ =0.00 %< 25% for heterogeneity between studies, suggesting no statistically significant between-study heterogeneity. The meta-analysis was applied using the fix-effects model.

#### 2.5.2 Meta-analysis of fixed effects

We meta-analysed the prevalence of clinical improvement from five studies. Based on the results of the meta-analyses, the pooled prevalence of clinical improvement significant findings was 62% (95% CI: 59-65%). Moreover, There was a statistically significant (Z=58.58, *p*=0.00<0.05). Table 3 shows the results of our meta-analyses for the prevalence of clinical improvement.

**Table 3.**
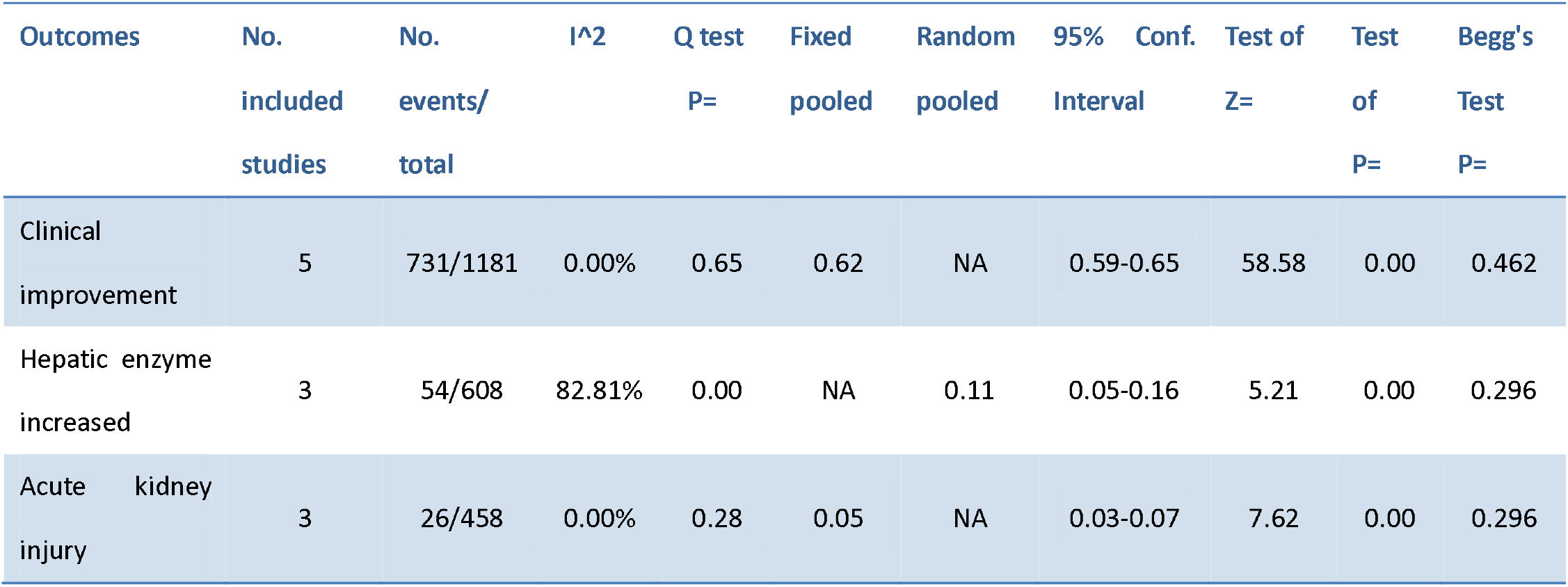
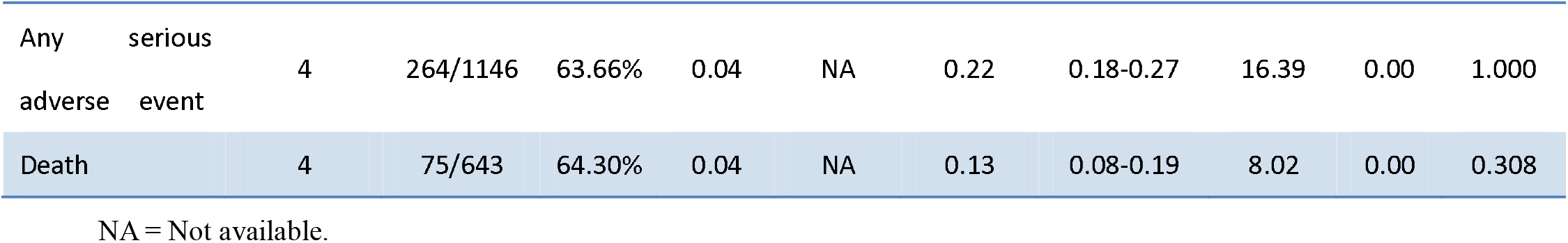
Single-Arm Study, Meta-analysis results. Table 3 shows the results of our meta-analyses for the prevalence of the clinical improvement, hepatic enzyme increased, acute kidney injury, any serious adverse event, and death.

#### 2.5.3 Bias test

Funnel plot and Begg’s test were used to evaluate potential publication bias. The shape of the funnel plot was almost symmetrical, suggesting that there was no publication bias. Furthermore, The results of Begg’s test (*p*=0.462) was indicated no evidence of statistically significant publication bias among the included studies.

### 2.6 Single-Ar m Study: Hepatic enzyme increased, meta-analysis results

#### 2.6.1 Heterogeneity test

For four studies [14, 16–18], Q-test *p*=0.00 < 0.1 and heterogeneity statistic *I*^*2*^ =91.4%> 75% for heterogeneity between studies, suggesting heterogeneity was fairly high. Sensitivity analysis was performed,to investigate the source of significant heterogeneity.

#### 2.6.2 Sensitivity analysis

Sensitivity analyses indicated that Antinori 2020 [14] study was the main origin of heterogeneity. After sensitivity analysis, we removed Antinori 2020 article, then *I*^*2*^ reduced to *I*^*2*^ =82.81%, *P*=0.00. However, the heterogeneity was already the lowest. After excluding Antinori 202 article, the meta-analysis was performed using random effects.

#### 2.6.3 Meta-analysis of random effects

We meta-analyzed the prevalence of Hepatic enzyme increased from three studies. Based on the results of the meta-analyses, the pooled prevalence of Hepatic enzyme increased, significant findings was 11% (95% CI: 5-16%). Moreover, There was a statistically significant (z=5.21, *p*<0.05). Table 3 shows the results of our meta-analyses for the prevalence of Hepatic enzyme increased.

#### 2.6.4 Bias test

Funnel plot and Begg’s test were used to evaluate potential publication bias. The shape of the funnel plot was almost symmetrical, suggesting that there was no publication bias. Furthermore, the results of Begg’s test (*p*=0.296) was indicated no evidence of statistically significant publication bias among the included studies.

### 2.7 Single-Arm Study: Acute kidney injury, meta-analysis results

#### 2.7.1 Heterogeneity test

In these four articles [14, 16–18], Q-test *p*=0.01<0.1 and heterogeneity statistic *I*^*2*^ =75.16%>75% for heterogeneity between studies, suggesting heterogeneity was fairly high. Sensitivity analysis was performed,to investigate the source of significant heterogeneity.

#### 2.7.2 Sensitivity analysis

Sensitivity analyses indicated that Wang 2020 [18] study was the main origin of heterogeneity. After sensitivity analysis, we removed Wang 2020 [18] article, then *I*^*2*^ reduced to *I*^*2*^ =0.00%, *p*=0.28, suggesting no statistically significant between-study heterogeneity. After excluding Wang 2020 [18] article, the meta-analysis was performed using fixed effects.

#### 2.7.3 Meta-analysis of fixed effects

We meta-analysed the prevalence of Acute kidney injury from three studies. Based on the results of the meta-analyses, the pooled prevalence of Acute kidney injury, significant findings was 5% (95% CI: 3-7%). Moreover, There was statistically significant (z = 7.62, *p* < 0.05). Table 3 shows the results of our meta-analyses for the prevalence of Acute kidney injury.

#### 2.7.4 Bias test

Funnel plot and Begg’s test were used to evaluate potential publication bias. The shape of the funnel plot was almost symmetrical, suggesting that there was no publication bias. Furthermore, the results of Begg’s test (*p*□= 0.296) was indicated no evidence of statistically significant publication bias among the included studies.

### 2.8 Single-Arm Study: Any serious adverse event, meta-analysis

#### 2.8.1 Heterogeneity test

In these five articles [14–18], Q-test *p* = 0.00 < 0.1 and heterogeneity statistic *I*^*2*^ =84.35%>75% for heterogeneity between studies, suggesting heterogeneity was fairly high. Sensitivity analysis was performed,to investigate the source of significant heterogeneity.

#### 2.8.2 Sensitivity analysis

Sensitivity analyses indicated that Antinori 200 [14] study was the main origin of heterogeneity. After sensitivity analysis, we removed Antinori 200 [14] article, then *I*^*2*^ reduced to *I*^*2*^ =63.66%, *p* = 0.04. Because the reason for heterogeneity could not be identified, after excluding Antinori 200 [14] article, the meta-analysis was performed using random effects.

#### 2.8.3 Meta-analysis of random effects

We meta-analyzed the prevalence of Any serious adverse event from four studies. Based on the results of the meta-analyses, the pooled prevalence of Any serious adverse event, significant findings was 22% (95% CI: 18-27%). Moreover, There was statistically significant (z = 16.39, *p* < 0.05). Table 3 shows the results of our meta-analyses for the prevalence of Any serious adverse event.

#### 2.8.4 Bias test

Funnel plot and Begg’s test were used to evaluate potential publication bias. The shape of the funnel plot was almost symmetrical, suggesting that there was no publication bias. Furthermore, The results of Begg’s test (*p*□=1.000) was indicated no evidence of statistically significant publication bias among the included studies.

### 2.9 Single-Arm Study: Death, meta-analysis results

#### 2.9.1 Heterogeneity test

In these five articles [14–18], Q-test *p*□= 0.00 < 0.1 and heterogeneity statistic *I*^*2*^ =80.21%>75% for heterogeneity between studies, suggesting heterogeneity was fairly high. Sensitivity analysis was performed,to investigate the source of significant heterogeneity.

#### 2.9.2 Sensitivity analysis

Sensitivity analyses indicated that Beigel 200 [15] study was the primary origin of heterogeneity. After sensitivity analysis, we removed Beigel 200 [15] article, then *I*^*2*^ reduced to *I*^*2*^ =64.30%, *p*□= 0.04. Because the reason for heterogeneity could not be identified, after excluding Beigel 200 [15] article, the meta-analysis was performed using random effects.

#### 2.9.3 Meta-analysis of random effects

We meta-analyzed the prevalence of Death event from four studies. Based on the results of the meta-analyses, the incidence rates of Death event significant findings was 13% (95% CI: 8-19%). Moreover, There was a statistically significant (z=8.02, *p*□< 0.05). Table 3 shows the results of our meta-analyses for the prevalence of Death event.

#### 2.9.4 Bias test

Funnel plot and Begg’s test were used to evaluate potential publication bias. The shape of the funnel plot was almost symmetrical, suggesting that there was no publication bias. Furthermore, The results of Begg’s test (*p*□=0.308) was indicated no evidence of statistically significant publication bias among the included studies.

## 3 Discussion

COVID-19 is an emerging infectious disease. It has been eight months so far, and there are no specific effects medicines for its treatment. The current clinical treatments for COVID-19 are directed mainly toward providing symptomatic management treatment [19]. Based on the in vitro studies of different coronaviruses (including SARS-CoV-2), it seems that many drugs may be candidate treatment options, including LPV/r, chloroquine, HCQ,remdesivir [12,20–25]. On May 1, 2020, after reviewing unpublished, available clinical trial data, the U.S. Food and Drug Administration issued an Emergency Use Authorization (EUA) to allow the use of remdesivir (a nucleotide analog), Inhibition of viral RNA-dependent RNA polymerase (RDRP), for severe coronavirus disease 2019 (COVID-19) in the treatment of adults and children hospitalized [26]. However, there is a lot of controversy about the efficacy and safety of remdesivir in the treatment of COVID-19[27–28]. Currently, in a randomized, double-blind, placebo-controlled study conducted in China, Wang et al [18] believed that remdesivir was not associated with statistically beneficial clinical outcomes for severe COVID-19. However, in another randomized placebo-controlled study, it showed that remdesivir has a beneficial effect on patients with severe COVID-19 [17]. In a placebo-controlled randomized controlled study of remdesivir, Antinori et al. [14] believed that remdesivir could benefit SARS-CoV-2 pneumonia patients hospitalized outside the intensive care unit (ICU). Therefore, it is important to provide evidence for the efficacy of remdesivir in the treatment of COVID-19.

In this systematic review and meta-analysis, based on two randomized, double-blind, placebo-controlled trials, the results showed that the clinical improvement of remdesivir in the treatment of COVID-19 was superior to the placebo-controlled group (Z=3.33, *p*□< 0.05). This indicates that remdesivir has a certain beneficial effect in the treatment of COVID-19, and Beigel et al. [15] believe that remdesivir is better than comfort in shortening the recovery time, and evidence of lower respiratory tract infection in adults hospitalized with Covid-19. In addition, in the one-arm study, it was suggested that the combination of remdesivir was significantly effective in treating COVID-19 (Z= 58.58, *p*□<0.05), and the effective rate was 62%. However, there is no placebo control in this study, and the currently included trials of remdesivir in the treatment of COVID-19 are all based on the combination of conventional treatment of COVID-19, so that, it can only show overall response rates of 62% in clinical improvement with remdesivir for COVID-19 patients. Antinori et al. [14],in a hospital-based control study between ICU patients and patients in infectious wards, remdesivir can benefit SARS-CoV-2 pneumonia patients who are hospitalized outside the ICU. Goldman et al [16], in a Based on a 5-day and 10-day treatment control study of remdesivir, found that there was no significant difference between the efficacy of the 5-day course and the 10-day course. Wang et al [18], argued that remdesivir is not associated with a statistically significant clinical benefit, but the amount of clinical improvement time was reduced in those who treated earlier. According to the Literature, a systematic review and meta-analysis of elevated liver transaminase, acute kidney injury, serious adverse events, and Death were conducted. Due to the lack of placebo-controlled studies, we conducted a single-arm study after data extraction. On the basis of the results of our meta-analysis including 3 studies with 608 patients,the results show that the incidence of Hepatic enzyme increased was 11%(z=5.21, *p* < 0.05), during patients received remdesivir treatment. In addition, We conducted a meta-analysis involving 3 studies with 458 participants to elucidate the Acute kidney injury, suggesting that the pooled prevalence of Acute kidney injury significant findings was 5% (z = 7.62, *p*□< 0.05), during patients received remdesivir treatment. Moreover, based on the results of our meta-analysis including 4 studies with 1,146 patients,the results show that the incidence of serious adverse events was 22%(z = 16.39, *p*□<0.05), during patients received remdesivir treatment. Based on the results above, this study showed that the incidence of elevated liver transaminase, acute kidney injury, and any serious adverse events in the treatment of COVID-19 with remdesivir is common. Due to the lack of a placebo-controlled, it is impossible to determine that directly lead to it due to the remdesivir side effects. However, attention should be paid. Antinori et al [14] believed that the most common adverse event during the treatment with remdesivir was liver toxicity, with elevated transaminases levels of 3-4 grade; the most common adverse event leading to treatment interruption was acute kidney injury. Beigel et al [15], based on a randomized, double-blind placebo-controlled study, believed that the most common adverse event in the remdesivir group was anemia or decreased hemoglobin ([7.9%], compared with the placebo group [9.0%]); acute kidney Injury events ([7.4%], and placebo group [7.3%]), but there was no significant difference in the incidence of adverse events between the remdesivir group and the placebo group. Grein et al [17] believe that the most common adverse events are elevated liver enzymes, diarrhea, rash, renal insufficiency, and hypotension. We conducted a meta-analysis involving 4 studies with 643 patients to elucidate mortality, suggesting that the hospital mortality was 13% (z=8.02, *p*□< 0.05), during patients received remdesivir treatment. Wang et al [18] believe that compared with placebo, the intravenous remdesivir of patients with severe COVID-19 cannot significantly improve the mortality rate or the time to clear the virus. The data on adverse effects and mortality with remdesivir, are the result of the meta-analysis of all five studies in various combinations, but, none of these results was demonstrated in the two large quality trials.

Based on our analysis, remdesivir is significantly effective in the treatment of COVID-19; besides, adverse events and mortality should also be paid attention. It is worthy of our further study. However, this meta-analysis has some limitations. First, fewer studies were included, especially double-blind placebo-controlled studies. Secondly, most of the indicators are single-armed studies and lack of comparison. The interpretation of these results is limited. Third, when combined with remdesivir in the treatment of COVID-19, it is affected by different drugs, such as other antiviral drugs. Finally, some studies have not been published.

## 4 Conclusion

This analysis confirms that remdesivir is effective in the clinical improvement of COVID-19 patients, and the rate of clinical improvement was 62%. We found that the incidence rates of Acute kidney injury, Hepatic enzyme increased, Any serious adverse event were 5%, 11%, 22%, respectively, and the mortality was 13%, during treatment of COVID-19 with remdesivir. However, the number of studies included analyses was limited, so more large-scale studies were needed to confirm the results, to further elucidate the underlying mechanisms.

## Data Availability

All data generated or analyzed during this study are included in this article.

## Abbreviations

COVID-19: Coronavirus disease 2019
RCT: Randomized controlled trial
RNA: Ribonucleic Acid

## Declarations

### Ethics approval and consent to participate

Not applicable. This article does not contain any studies with human participants performed by any of the authors.

## Consent for publication

The authors have obtained the patient’s written informed consent for publication of this case report and any accompanying images. The proof of consent to publish from the patient can be requested at any time.

## Availability of data and materials

All data generated or analyzed during this study are included in this article.

## Competing interests

The authors declare that they have no competing interests.

## Funding

The authors state that this work has not received financial support.

## Authors’ contributions

All the authors designed the study. Lixiang Lou and Hui Zhang Li designed the literature search and searched the articles. Zeqing Li and Baoming Tang contributed to collect patient data. Lixiang Lou and Hui Zhang wrote the first draft of the article. All the authors revised the article and approved the final version.

## Acknowledgements

Not applicable.

## Notes

### Competing Interest Statement

The authors have declared no competing interest.

